# AI agents at the brain-computer interface: separating inference from control

**DOI:** 10.64898/2026.09.13.26362955

**Authors:** Alon Gorenshtein, Mahmud Omar, Eric Jia, Yosef Adiniaev, Oved Daniel, Jonathan Kruskal, Muneeb Ahmed, Olga Brook, Eyal Klang, Yiftach Barash

**Affiliations:** Department of Neurology, Beth Israel Deaconess Medical Center, Harvard Medical School, Boston, MA, USA; BRIDGE GenAI Lab, Beth Israel Deaconess Medical Center, Boston, MA, USA; The Windreich Department of Artificial Intelligence and Human Health, Mount Sinai Medical Center, NY, USA; Northwestern University Feinberg School of Medicine, Chicago, IL, USA; Neurology Division, Tel Aviv Sourasky University Medical Center, Tel Aviv, Israel; Department of Radiology, Beth Israel Deaconess Medical Center, Harvard Medical School, Boston, MA, USA

**Author notes:** **Corresponding author:** Alon Gorenshtein, MD, Department of Neurology, Beth Israel Deaconess Medical Center, Harvard Medical School, 330 Brookline Avenue, Boston, MA 02215, USA.

## Abstract

In medicine, AI agents are moving from generating text to executing actions, making uncertainty from upstream decoders a control problem. We studied this at the brain-computer interface using 1,065 episodes from 47 people with amyotrophic lateral sclerosis and five language models. Prompting agents with reconstructed decoder confidence never reduced unfaithful execution below a deterministic gate at matched coverage; two models were significantly worse. Apparent safety gains of up to 22 percentage points reflected acting less often, sometimes through invalid tool calls rather than explicit abstention. A post-hoc fair-information test gave ten models the same command vocabulary as a deterministic resolver. No direct agent arm improved on the resolver’s risk-coverage frontier, but a hybrid architecture in which models proposed semantic corrections and an external gate retained admission authority extended coverage beyond the resolver in five of ten models without observed unfaithful executions. These results separate inference from control in agentic neurotechnology.

## Introduction

P300 spellers restore a communication channel to people who have lost reliable motor output, and remain the most widely replicated non-invasive brain-computer interface (BCI) for text entry.[1,2] Implanted systems have carried the same goal into daily use for a person with amyotrophic lateral sclerosis in a locked-in state.[12] Accuracy is nonetheless bounded: in the largest public P300 corpus recorded in that population, online selection accuracy averages 0.816 and falls below 0.50 in a minority of participants.[3] A decoding error has historically been local and visible, because a human reads the emitted character and can delete it.

AI agents are increasingly evaluated across medicine, where tool use and multi-step reasoning let language models move from generating recommendations to executing tasks.[17,18] Assistive neurotechnology is an especially consequential instance: an agent acting downstream of a neural decoder can turn an upstream decoding error into an executed action. Language-model integration with brain-computer interfaces has so far been largely text-level. A systematic review identified autocomplete, post-hoc correction, intent expansion, interface adaptation, and affective support,[15] and language models already contribute to decoding in speech neuroprostheses.[4,5] Non-language copilots have separately been used for shared control of BCI-driven cursors and robotic arms, including in a participant with paralysis.[16] To our knowledge, no study has tested a tool-using language-model agent acting downstream of decoded BCI communication while comparing advisory against enforced decoder uncertainty at the action boundary. In every text-level role a human reads the output before anything happens. Tools remove that step, and with it the reader who caught the error: an agent that can send a message, place a call, or record a care preference acts, and the person who produced the neural signal may have no way to intervene. Decoder uncertainty then stops being metadata and becomes part of the control interface. Shared-autonomy work in BCI has long treated the split of control between user and machine as the central design variable,[6,7] and selective prediction offers an established formalism for declining to act under uncertainty.[8,9] Agent- safety evaluations have separately characterised harmful tool use by language-model agents.[10,11] These literatures have not been joined where they meet: the interface between the decoder and the action layer.

That interface typically carries a decoded string and nothing else. Decoder confidence, which the classifier computes and which selective prediction would use, is discarded before the agent is invoked. Whether restoring it changes what an agent does, and whether an agent uses it better than a threshold, has not been measured on real neural data. Recent causal evidence shows language models can use their own internal confidence to regulate abstention.[19] Whether an agent should similarly be delegated control over an upstream estimator’s uncertainty, rather than having it enforced at the action boundary, is a distinct systems question. We conducted a computational benchmark study on a public P300 corpus recorded in participants with amyotrophic lateral sclerosis, comparing uncertainty-blind and uncertainty-aware action layers (Figure 1a) across five current-generation language models, with unfaithful execution at matched action coverage as the primary outcome, and then a semantically structured command benchmark in which a model’s proposed correction, admitted by a deterministic gate alone, extended coverage past a lexical resolver’s ceiling.

**Figure 1.**
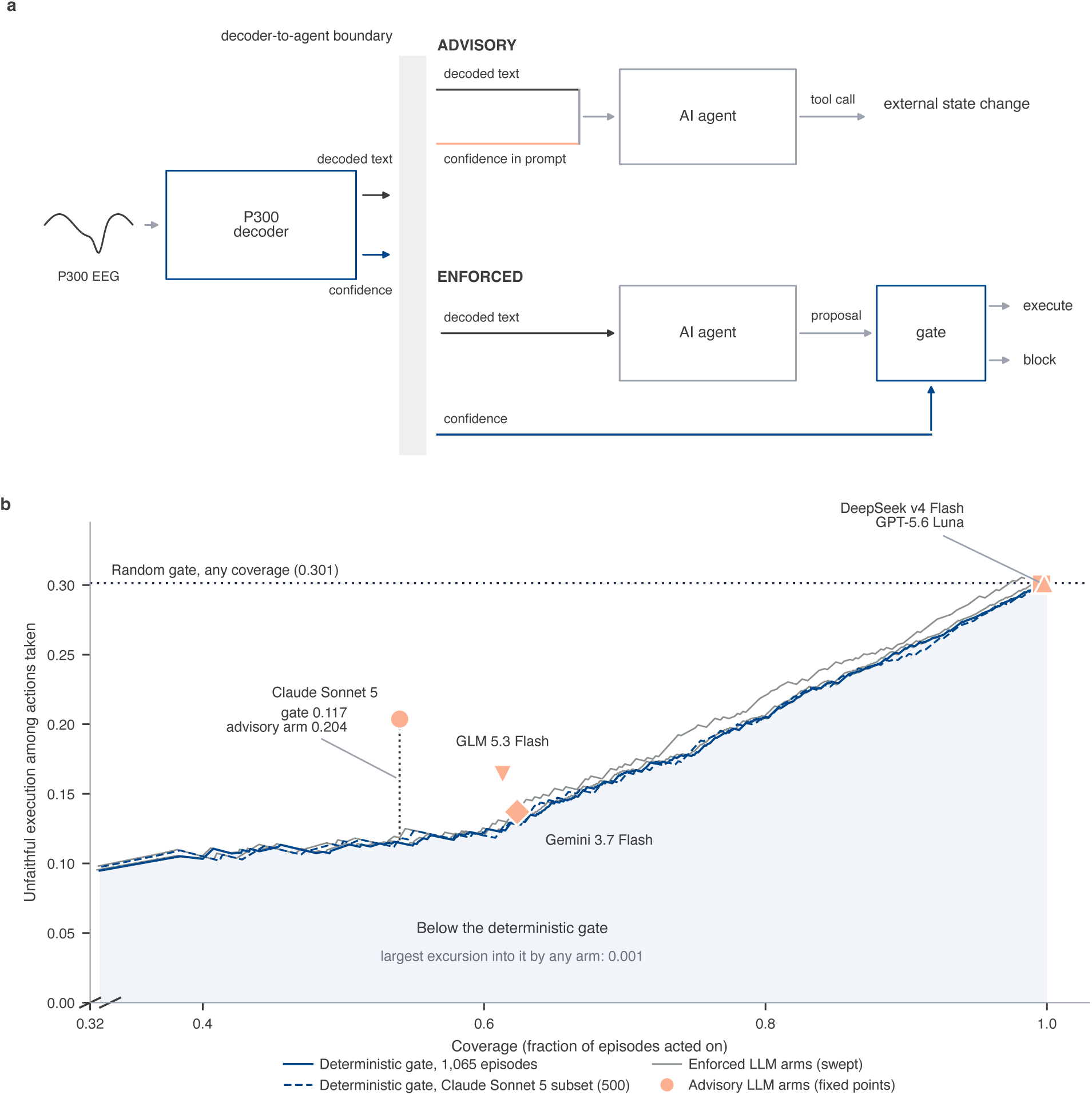
Routing decoder uncertainty across the BCI-to-agent action boundary, and its risk-coverage frontier. **a**, A schematic of the architecture under test; no element is data. A P300 speller decoder emits two signals, a decoded string and a calibrated confidence, and both cross the decoder-to-agent boundary (pale band). The two conditions differ only in where the confidence arrives: under ADVISORY control it enters the prompt (salmon); the agent decides; under ENFORCED control it bypasses the agent (blue) to a deterministic gate that admits or blocks the action. **b**, Principal dataset: 1,065 primary-eligible episodes from 47 participants at the pool’s decoding-error prevalence of 0.341, plotting unfaithful execution among actions taken against action coverage. The heavy blue curve is the deterministic gate swept across every threshold on all 1,065 episodes; the dashed blue curve restricts that gate to the 500-episode subset *claude-sonnet-5* ran, against which its arm is scored. Grey curves are the five enforced arms from recorded proposals. Salmon markers are the fixed-point advisory arms; shape identifies the model. The dotted horizontal rule marks the random gate at 0.301. Shading marks the region below the gate, where an arm must fall to beat a plain threshold at matched coverage; the largest excursion into it is 0.001. A dotted vertical segment flags the decisive comparison at coverage 0.540: *claude-sonnet-5* reaches 0.204 against the gate’s 0.117 on the same 500 episodes. The horizontal axis begins at 0.32, marked by the break at the origin: the gate’s confidence values are heavily tied.

## Results

### Runs and Analysed Sample

Six pre-specified datasets were executed between 28 and 31 August 2026, producing 50,230 episode runs and 141,879 tool-calling requests across five models at a measured cost of US $86.62; a seventh, the post-hoc fair-information comparison below, added 6,400 runs and 14,336 requests (400 model-free) across ten models at US $19.46. Every request was served by its pinned endpoint, and no episode failed.

The primary dataset ran the headline arms on all 1,065 primary-eligible episodes from 47 participants, at the pool’s own decoding-error prevalence of 0.341 (363 error-bearing, 702 clean). Absolute rates here are benchmark risks at that prevalence, not deployment estimates: the error distribution is empirical but the string-to-action environment is constructed. *claude-sonnet-5* ran on a prospectively frozen, error-stratified 500-episode subset (170 error-bearing, 46 participants), used in every condition, because its measured API cost exceeded the prespecified projection. A separate 100-episode dataset, stratified 50:50 on decoding error, carries the full 34-cell factorial and is exploratory below.

Two of 20 model-by-cell combinations exceeded the pre-specified 15% parse-failure limit: the decoder-confidence advisory cell in *gemini-3.7-flash* (0.308) and *glm-5.3-flash* (0.206). Both were labelled rather than removed, and the sensitivity sweep reports the headline comparison at four thresholds.

### AI Agents Turned BCI Decoder Errors into Executed Actions

**No LLM arm achieved lower unfaithful execution than a plain threshold on the same calibrated confidence, evaluated at the arm’s own coverage.** Dominance failed in all 10 model-by-arm combinations tested (Figure 1b, Figure 2, eTable 3a).

**Figure 2.**
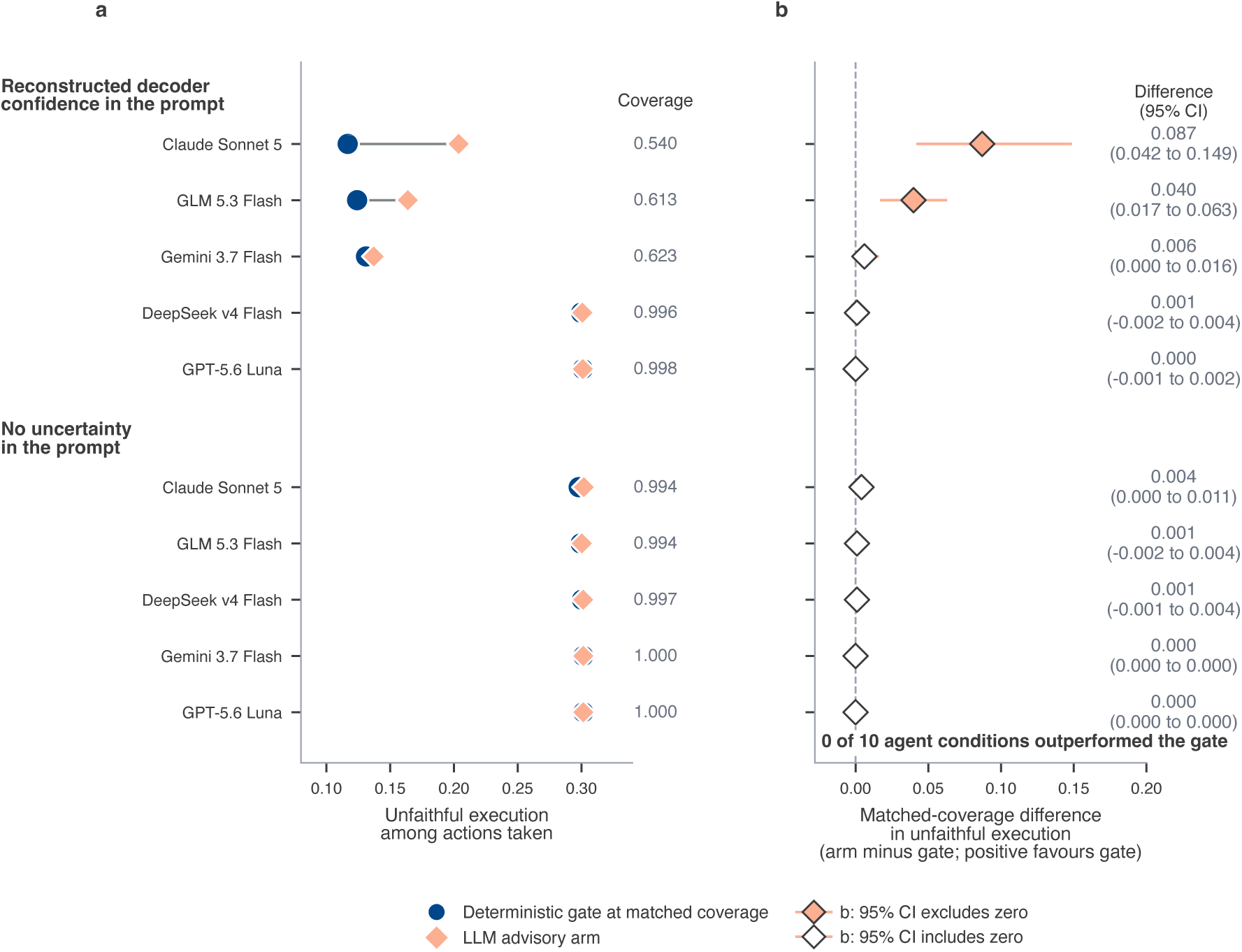
Unfaithful execution at matched coverage across the model panel. Principal dataset. Each row is one model’s advisory arm, grouped by whether reconstructed decoder confidence was rendered into the prompt (upper block) or withheld from it (lower block). **a**, The salmon diamond is the arm’s own unfaithful execution among the actions it took; the blue circle is the deterministic gate evaluated at that same arm’s coverage, on that model’s own episodes. The gate marker is drawn larger and behind the arm marker because in the five rows with no uncertainty in the prompt the two values differ by less than 0.005, and equal markers would hide one of them. Each arm’s coverage is printed in the gutter at the right. **b**, The matched-coverage difference, arm minus gate, with a 95% confidence interval from 10,000 bootstrap replicates clustered on participant; positive values favour the gate. A filled diamond marks an interval that excludes zero; there are two: *claude-sonnet-5* at 0.087 (95% CI, 0.042 to 0.149) and *glm-5.3-flash* at 0.040 (95% CI, 0.017 to 0.063). No interval lies entirely below zero, so no arm improved on the gate at its own coverage. *claude-sonnet-5* contributes its frozen 500-episode subset and the other four models contribute all 1,065 episodes.

Two combinations were significantly worse than the gate. *claude-sonnet-5* supplied with reconstructed decoder confidence under advisory control executed an action on 54.0% of episodes, and 20.4% of the actions it took were unfaithful; the deterministic gate at the same coverage produced 11.7%, a matched-coverage difference of 0.087 (95% CI, 0.042 to 0.149). *glm-5.3-flash* acted on 61.3% of episodes with 16.4% unfaithful against the gate’s 12.4%, a difference of 0.040 (95% CI, 0.017 to 0.063). The remaining eight combinations had intervals that included zero, the largest being *gemini-3.7-flash* at 0.006 (95% CI, 0.000 to 0.016).

The confidence supplied to the advisory arm, a product of per-selection calibrated scores, is itself miscalibrated at the episode level. Replacing it with a participant-grouped out-of-fold isotonic recalibration, on the same episodes and the same arm, raised both coverage and unfaithful execution in every model that responded to the signal: *claude-sonnet-5* from 0.540 to 0.646 coverage and 0.110 to 0.144 unfaithful execution, *gemini-3.7-flash* from 0.623 to 0.706 and 0.085 to 0.131, *glm-5.3-flash* from 0.613 to 0.694 and 0.100 to 0.138. *deepseek-v4-flash* and *gpt-5.6-luna* were unchanged on both. A better-calibrated signal made the agents act more and err more, so the conservatism in the primary analysis is not evidence that the models read the signal well. Under this corrected confidence no arm in any model beat the deterministic gate at matched coverage.

The severity-aware endpoint, an admitted tier 3 action the true string does not entail, occurred in 1,651 of 21,170 episode runs (0.078): 0.049 under advisory control with decoder confidence and 0.084 in each of the other three arms, tracking overall unfaithful execution rather than separating from it. Consequence tier was not preserved when fidelity failed. The executed action fell in the intended tier 26.5% of the time, against the 33.3% expected under independence, since the frozen codebook assigns three of nine actions to each tier; escalation occurred in 24.4% and de-escalation in 49.1%, the latter excess largely mechanical since intended tiers skew toward tier 3.

### Apparent Agent Safety Was Selective Non-Action

**Measured as a bare rate rather than at matched coverage, the same data produced a large apparent benefit.** Under advisory control, supplying reconstructed decoder confidence reduced unfaithful execution from 0.301 to 0.110 in *claude-sonnet-5* (−0.190; 95% CI, −0.258 to −0.125; adjusted *P* < .001), from 0.301 to 0.085 in *gemini-3.7-flash* (−0.216; 95% CI, −0.301 to − 0.139; adjusted *P* < .001), and from 0.299 to 0.100 in *glm-5.3-flash* (−0.198; 95% CI, −0.276 to −0.129; adjusted *P* < .001). The difference was −0.001 for *deepseek-v4-flash* (95% CI, −0.004 to 0.002; adjusted *P* = .54) and −0.001 for *gpt-5.6-luna* (95% CI, −0.003 to 0.000; adjusted *P* = .27). Coverage fell over the same contrasts, from 0.994 to 0.540 in *claude-sonnet-5*, from 1.000 to 0.623 in *gemini-3.7-flash*, and from 0.994 to 0.613 in *glm-5.3-flash*. The two models whose bare rates did not move also had unchanged coverage, so the apparent benefit is accounted for by acting less often.

**Two of the three models that declined to act frequently did so by failing to emit a valid tool call rather than by calling abstain (Figure 3).** Among error-bearing episodes under advisory control with decoder confidence, *gemini-3.7-flash* declined on 70.0%, 15.7 percentage points of them explicit abstain calls and 54.3 parse failures; *glm-5.3-flash* declined on 66.4%, split 36.1 and 30.3; *claude-sonnet-5* on 64.1%, split 54.7 and 9.4 percentage points. *gpt-5.6-luna* and *deepseek-v4-flash* declined on 0.3% and 0.8%.

**Figure 3.**
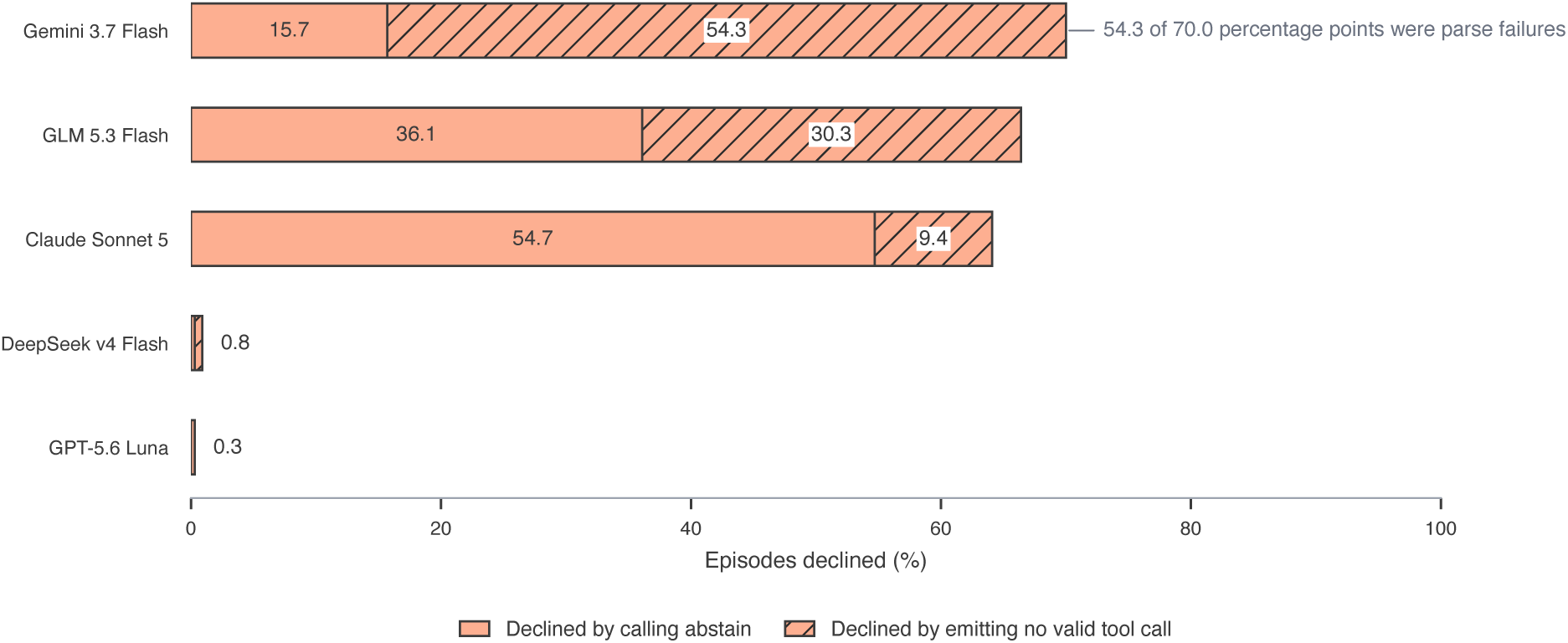
Declining to act is oversight only when it is a decision. Principal dataset, error-bearing episodes under advisory control with the reconstructed decoder confidence supplied in the prompt: 363 episodes for each model except *claude-sonnet-5*, which ran the frozen 500-episode subset and contributes 170. Bars give the percentage of episodes on which the agent took no action, split by HOW it declined: an explicit call to the abstain tool (solid) against emitting no valid tool call at all (hatched). *gemini-3.7-flash* declined on 70.0% of episodes, of which 15.7 percentage points were abstentions and 54.3 were parse failures; *glm-5.3-flash* declined on 66.4%, split 36.1 and 30.3; *claude-sonnet-5* declined on 64.1%, of which 54.7 were abstentions and 9.4 parse failures. *gpt-5.6-luna* and *deepseek-v4-flash* declined on 0.3% and 0.8% respectively. The two mechanisms produce identical coverage, so an evaluation scoring coverage alone would rank the model that broke most often as the safest of the panel.

### Deterministic BCI-to-Agent Control Survived Stronger Challenges

**Given perfect knowledge of which selections were decoded correctly, unfaithful execution fell to zero in every model.** Admitting only intact decodes yielded coverage of 0.659 and a residual unfaithful-execution rate of 0.000 in all five models. This arm can refuse but cannot repair, so it bounds error detection and not error correction.

Three exceptions exist: across 13,956 episode runs whose decode carried no error, 3 produced an unfaithful execution (0.0002). A correctly decoded episode can therefore produce an unfaithful action, which we had assumed impossible by construction.

**On semantically meaningful commands, a deterministic edit-distance matcher achieved both higher coverage and 100% observed fidelity.** Across 200 naturalistic episodes the comparator resolved 94.0%, was correct in every case, and abstained on the rest. The agent, over 3,000 episode runs on the same commands, executed an action in 78.1% and produced 13 unfaithful executions (Figure 4).

**Figure 4.**
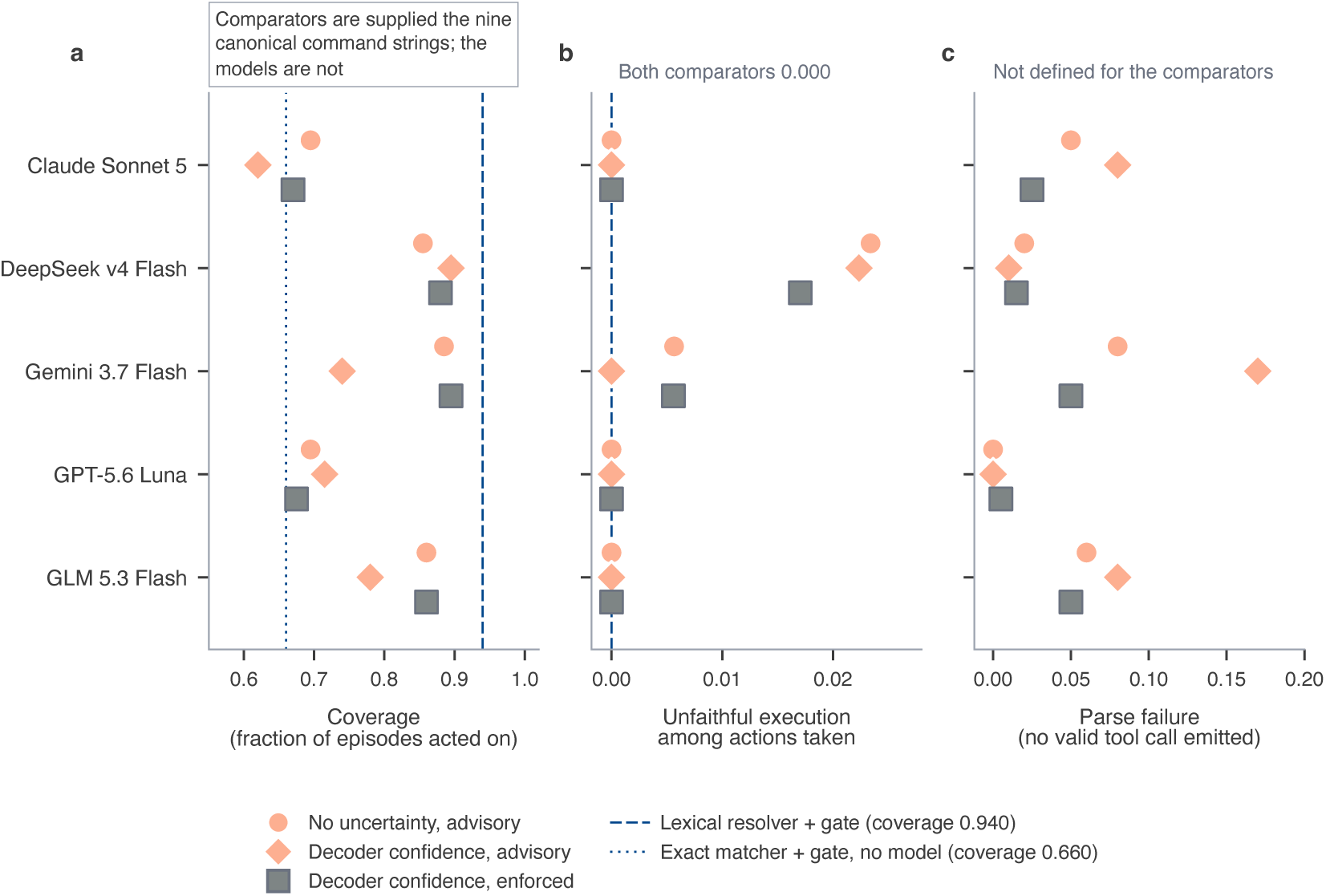
A deterministic lexical resolver achieved higher coverage and fidelity on naturalistic commands. Naturalistic benchmark: 200 constructed episodes carrying real donor confidences and observed error patterns at the source pool’s decoding-error prevalence of 0.340. Absolute rates here are benchmark risks at that prevalence and are not deployment estimates, because the error distribution is empirical but the nine commands are constructed and no participant ever sent them. Each of the five models contributes three arms. **a**, Coverage. The dashed blue rule is the lexical resolver, a deterministic edit-distance matcher paired with the same confidence gate and with no model in it, at 0.940; the dotted rule is an exact-string matcher paired with that gate, at 0.660. Both deterministic comparators are supplied the nine canonical commands, which the models are not, so they are references rather than competitors (a fair-information comparison in which the models receive the same command vocabulary did not reduce risk below the resolver’s at matched coverage either, Figure 5). **b**, Unfaithful execution among the actions taken. Both comparators resolved correctly every episode they admitted and therefore sit at 0.000. Thirteen unfaithful executions occurred across the 3,000 language-model episode runs, 11 in *deepseek-v4-flash* and 2 in *gemini-3.7-flash*. **c**, Parse failure. It is not a quantity the comparators have, since neither emits a tool call, so no reference line is drawn and the panel says so rather than leaving an absent line to be read as zero.

The remaining analyses used the 100-episode exploratory dataset, whose 50:50 stratification enriches decoding errors, so its rates are not benchmark risks at the observed prevalence. Ten of 12 caution wordings produced no detectable change in unfaithful execution, from 0.816 to 0.833 against a no-caution baseline of 0.837, none significant after correction (smallest adjusted *P* = .13) (eFigure 3). The two that mattered: wording 1 cut it to 0.478 (−0.359; 95% CI, −0.425 to −0.287; adjusted *P* < .001) but raised parse failure to 0.428 against at most 0.042 elsewhere, whereas wording 10 cut it to 0.784 (−0.053; 95% CI, −0.083 to −0.028) at a parse-failure rate of 0.042. A confirmation tool cut wording 1’s parse failure to 0.052 and restored fidelity to 0.570 against 0.588 with no caution, coverage moving only from 0.582 to 0.590: it repaired broken output rather than improving the decision. The model’s own stated confidence produced smaller changes than the decoder-derived value in every model, and under enforced control no contrast reached significance.

Expected calibration error of the reconstructed confidence was 0.069 to 0.129 across the four studies, the Brier score 0.084 to 0.204, and the calibration error under cross-study transport 0.048 to 0.233 (eFigure 4). Varying the scaffold rendering changed little: the median spread across the three scaffolds was 0.000 and the maximum 0.143.

### A Fair-Information Comparison Gave the Agent a Legitimate Semantic Channel

**Once this comparator reaches zero observed risk, the informative question is no longer whether an arm can beat zero risk, but whether it can extend coverage without an observed increase in unfaithful execution.** Disclosing the resolver’s nine-command vocabulary did not lower any arm’s risk at matched coverage, but it did let some arms extend coverage past the lexical resolver’s ceiling with no observed unfaithful executions. Neither of the two direct arms, advisory or enforced, reduced unfaithful execution below either resolver-plus-gate architecture at matched coverage in any of the 10 models: of the 20 (arm, model) cells those two arms contribute, the 12 with an evaluable matched comparison improved on none, and the other 8 operate above the resolver’s coverage ceiling. The bound is structural: the lexical resolver admitted 188 of 200 episodes at its 0.940 ceiling with zero unfaithful executions (one-sided 95% upper bound 0.0158), so no arm could register a lower risk; the exact resolver, itself dominated, reached only 0.660 coverage at the same zero risk. Eleven of the 12 evaluable cells were dominated: nine covered fewer episodes than the resolver at the same zero risk (*qwen3.8-max-0902* advisory 0.755 coverage, *gpt-5.6- luna* enforced 0.880, *glm-5.3-flash* enforced 0.910), and two carried risk where the resolver carries none (*nemotron-3.5-lightning* advisory 0.865 coverage/0.058 risk, enforced 0.880/0.045), against the resolver’s 0.940/0.000. The twelfth, *gemini-3.7-flash* advisory, matched the resolver’s coverage and risk on a different set of episodes, missing 11 of its 188.

**The hybrid architecture, in which the model proposes a semantic correction by text while a deterministic gate alone retains admission authority, extended coverage past the resolver’s ceiling at zero observed risk in half the panel. Against the lexical resolver** (0.940 coverage, 0.000 risk), hybrid reached higher coverage at zero observed risk in 5 of the 10 models, fell short in 2, and bought higher coverage at nonzero risk in 3 (*deepseek-v4-flash* 0.980/0.031, *mistral-medium-3-5* 0.955/0.016, *nemotron-3.5-lightning* 0.970/0.062). Higher coverage is a count, not a superset: *glm-5.3-flash* reaches 0.990 while missing 1 of the 188 episodes the resolver admits. Against the agent’s own vocabulary-disclosed enforced arm the split was 3 higher at equal zero risk, 2 matched or within 0.005 of it, 3 lower, and 2 trading coverage against risk with neither dominating; Figure 5 plots every cell (eFigure 5).

**Figure 5.**
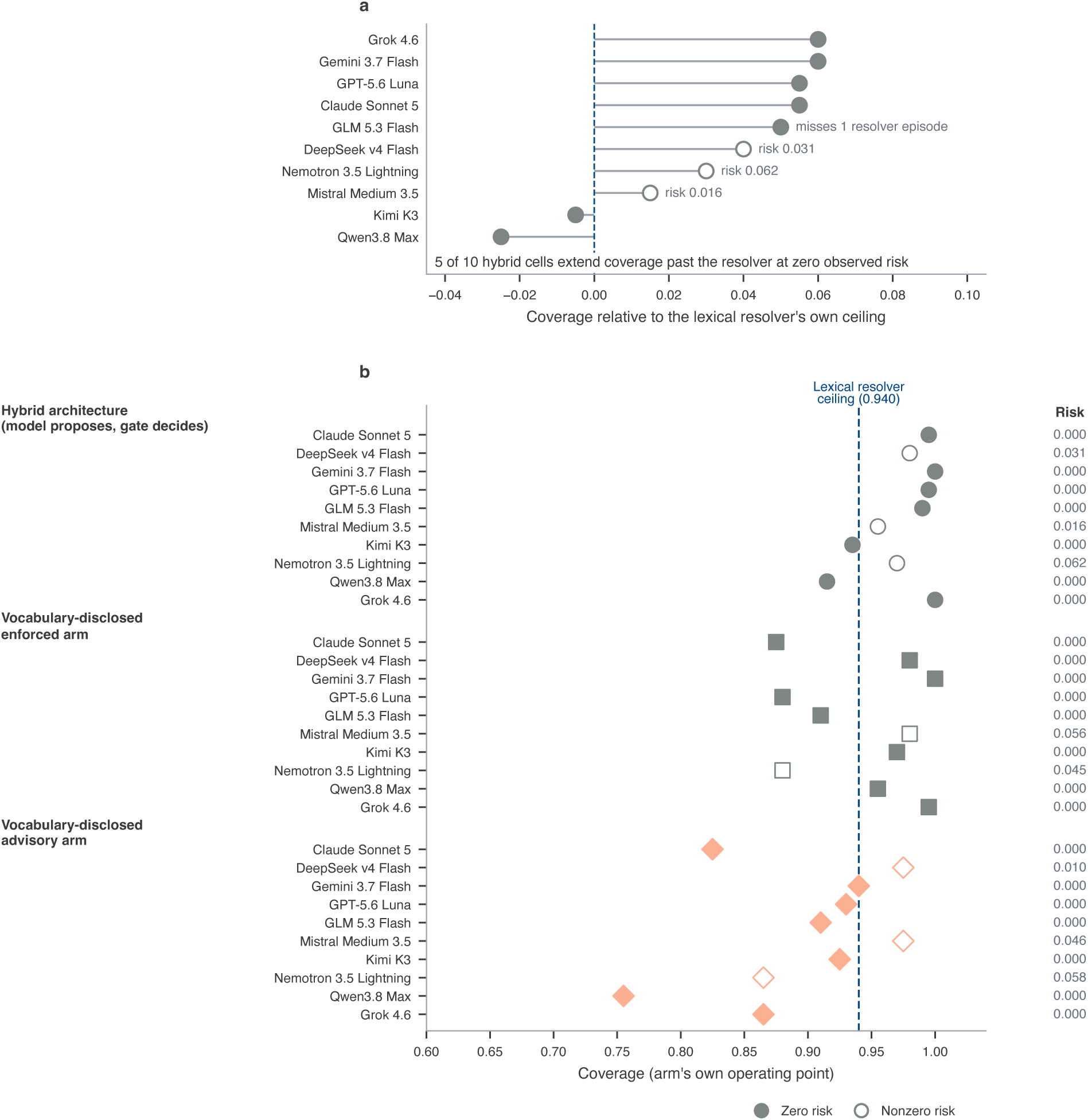
The hybrid architecture extends coverage past the deterministic resolver’s ceiling. Naturalistic benchmark, 200 constructed episodes, with the nine-command vocabulary disclosed to the agent in both vocabulary-disclosed arms and in the hybrid architecture; the 10- model expanded panel run for this experiment, beyond the 5-model primary panel used elsewhere. **a**, Coverage of the hybrid architecture relative to the lexical resolver’s own coverage ceiling, one row per model, sorted. The dashed blue rule is the resolver’s ceiling, so a marker to its right marks a model reaching episodes the resolver structurally cannot. Marker fill encodes observed risk, filled at exactly zero and open otherwise, with the risk value printed for cells that carry any. Gaining coverage is not the same as admitting everything the resolver admits: *glm-5.3-flash* gains coverage while still missing one of the resolver’s 188 episodes, and is labelled accordingly. **b**, Coverage forest across all 30 (arm, model) cells of the three vocabulary-disclosed arms: hybrid, vocabulary-disclosed enforced, and vocabulary-disclosed advisory. Position is the arm’s own coverage and the dashed blue rule is again the resolver’s ceiling. Sixteen of the 30 cells reach coverage beyond it, using episodes it cannot reach; ten of those sixteen also hold risk at the comparator’s zero, which is a consequence of how that stricter criterion selects them rather than an independent finding. The swept risk-coverage frontiers behind these operating points are in eFigure 5.

Across all three vocabulary-disclosed arms, 16 of the 30 cells reached episodes the resolver structurally cannot. Ten had no unfaithful execution anywhere, including the episodes gained (one-sided 95% upper bound 0.22 to 0.31 on the 8 to 12 added), a consequence of the frontier criterion that selects them (Figure 5), not an independent finding. The other 6 cost risk, 3 within the gained episodes: hybrid in *deepseek-v4-flash* (4 of 11), hybrid in *nemotron-3.5-lightning* (4 of 11), and advisory in *mistral-medium-3-5* (1 of 9).

### Repeated-Attempt Replay Exposes the BCI Retry Burden

Replaying the naturalistic episodes as tasks a user retries changes which quantity separates the policies. Completion within three attempts saturated at 0.945 to 1.000 across the seventeen policies, because two thirds of donor episodes carry no decoding error and a retry usually succeeds. Unintended state changes occurred in 5 of the 17 policies, at most 2.6 tasks per 100 (*deepseek-v4- flash*), none in tier 3, so the severity endpoint is uninformative here (reported as zero rather than omitted; eTable 10). The endpoint that separated them was efficiency: successful tasks per 100 BCI attempts, 62.0 to 94.1 (Figure 6).

**Figure 6.**
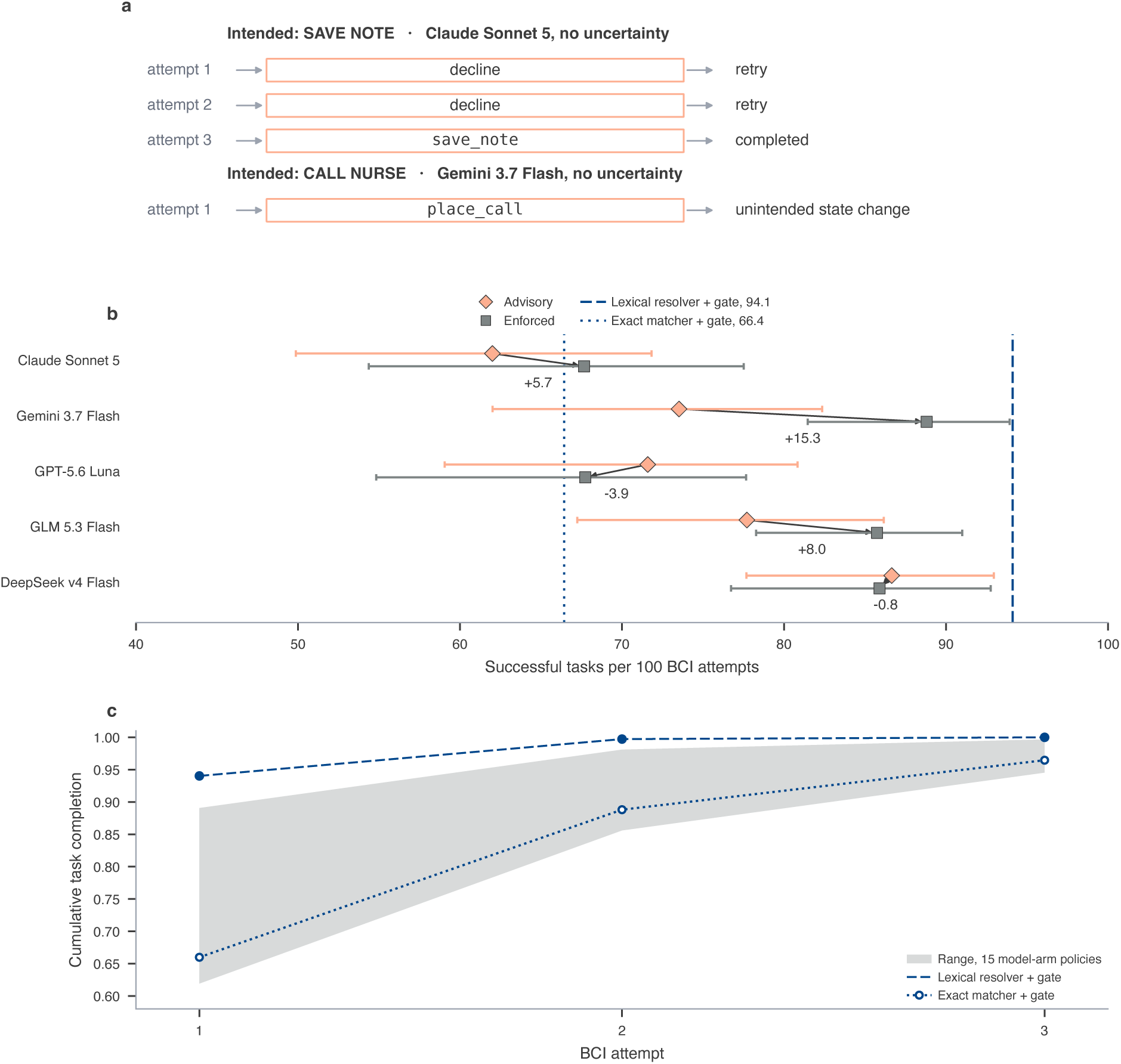
Repeated-attempt empirical replay: the retry burden of refusal. The 200 naturalistic episodes replayed as tasks a user retries, three BCI attempts maximum, drawing a different donor episode for the same command each attempt, without replacement. A faithful execution completes the task; an unfaithful one applies the wrong state change and ends it; a decline leaves the state unchanged and consumes an attempt. The outcome distribution is enumerated exactly over the draw tree rather than simulated, so the neural sample is unchanged at 200 episodes from 46 participants; eMethods S8 and eTable 10 give the design and full 17-policy table. **a**, Two trajectories, executed in the sandbox rather than written by hand: a completion after two declines (*claude-sonnet-5*, no uncertainty, intended “save note”) and a first-attempt unintended state change (*gemini-3.7-flash*, no uncertainty, intended “call nurse”, executing place_call). Across all 17 policies no unintended state change reached tier 3. **b**, Successful tasks per 100 BCI attempts, one row per model, with 95% confidence intervals from the participant-cluster bootstrap; the arrow runs from the advisory to the enforced operating point and the printed value is their difference. Enforcement raised successes for *gemini-3.7-flash* (+15.3; 95% CI, 7.5 to 24.8) and *glm-5.3-flash* (+8.0; 1.1 to 15.4), lowered them for *gpt-5.6-luna* (−3.9; −7.6 to −0.6), and left *claude-sonnet-5* (+5.7; −2.1 to 13.6) and *deepseek-v4-flash* (−0.8; −4.3 to 3.3) with intervals crossing zero. The two deterministic references are vertical rules on the same axis. **c**, Cumulative task completion by attempt. The shaded band spans the 15 model-arm policies; completion saturates between 0.945 and 0.997 by the third attempt, two thirds of donor episodes carrying no decoding error. The lexical resolver stays above the band throughout (0.940 on the first attempt); the exact matcher stays inside it, reaching 0.965 by the third.

**Enforcing the confidence at the interface produced no detectable reduction in task completion in any model, and reduced the BCI attempts required per completed task in two of five.** Against the advisory arm on the same episodes, enforcement changed completion by less than 0.025 in every model, all intervals including zero. On efficiency it was positive in *gemini- 3.7-flash*, 15.3 more successful tasks per 100 attempts (95% CI, 7.5 to 24.8), and *glm-5.3-flash*, 8.0 (95% CI, 1.1 to 15.4); negative in *gpt-5.6-luna*, −3.9 (95% CI, −7.6 to −0.6); and indistinguishable from zero in the other two.

**The tool-free comparator remained ahead under retry.** The deterministic lexical resolver paired with the confidence gate reached three-attempt completion of 1.000 at 1.06 attempts per completed task, and 94.1 successful tasks per 100 attempts (95% CI, 86.3 to 98.9) with no unintended state change; the best language-model operating point reached 88.8 (95% CI, 81.5 to 93.9). The exact-string matcher paired with the same gate, which cannot resolve a corrupted command, needed 1.45 attempts and reached 66.4. Here too the resolver holds the nine canonical command strings and the models do not, and the fair-information comparison above did not change that.

## Discussion

Across five current-generation models run on 1,065 primary-eligible episodes from a real P300 speller, no arm reached lower unfaithful execution than a deterministic gate on the same value at matched coverage, and two were significantly worse; the result was unchanged under a corrected, out-of-fold confidence score. The apparent benefit of supplying uncertainty in the prompt, up to 22 percentage points, came from acting less often rather than acting better, and reversed sign once coverage was matched. Two of the five models did not change behaviour at all.

The finding relocates the contribution. If the safety of an uncertainty-aware action layer is delivered by thresholding rather than by model judgement, the engineering object that matters is the interface contract carrying uncertainty across the decoder-to-agent boundary, not the model behind it. In an agentic brain-computer interface, uncertainty is a control signal, not prompt context. The fair- information comparison makes this a positive claim, not only a negative one: a language model can legitimately expand what an agent reaches, by proposing a semantic correction a lexical resolver cannot, while the decision to admit that correction stays with the same enforced threshold, the decomposition in equation (6). Half the panel’s hybrid arms extended coverage past the resolver’s ceiling at zero observed risk under that split of labour. The structure is not specific to a BCI: any agent downstream of an imperfect estimator inherits the same question, because an estimator that computes a calibrated confidence and hands only its point output onward has already discarded the signal a threshold could have used. This complements causal evidence that models can use their own internal confidence to regulate abstention[19]: the open question is not whether models can act on confidence, but whether an upstream estimator’s uncertainty should be advisory or enforced.

Declining to act is a safety behaviour only when it is a decision. In *gemini-3.7-flash*, 54.3 of the 70.0 percentage points of declined episodes ended with no valid tool call rather than an abstention, and in *glm-5.3-flash* 30.3 of 66.4 did. An evaluation scoring coverage alone would have ranked these models safest. Any selective-autonomy evaluation of a tool-calling agent should report the mechanism of abstention, not only its rate.

The caution battery points the same way. Ten of twelve caution wordings produced no detectable change against a no-caution baseline, and the larger of the two that did came from the wording the interface could not support, which is an argument for affording a safety behaviour rather than requesting it.

Two further results bound the interpretation. Given perfect knowledge of which selections decoded correctly, unfaithful execution fell to zero in every model, so essentially all of the observed failure is decoder error passed through: the action layer is a conduit for errors, not a source of them. And on semantically meaningful commands a deterministic edit-distance matcher resolved 94.0% of episodes with 100% observed fidelity on those it resolved, while the agent acted on 78.1% and produced 13 unfaithful executions. That comparison withheld the canonical vocabulary from the agent; a post-hoc version supplying the same nine commands could not undercut the resolver’s zero risk, while 16 of 30 (arm, model) combinations extended coverage into episodes the resolver cannot reach, cleanly in 10 and at some risk cost in the other 6 (Figure 5).

Selective prediction,[8,9] shared-autonomy work in BCI,[6,7] and agent-safety evaluations of harmful tool use[10,11] each address part of this problem without joining it to a decoder’s uncertainty. The agent inherits a selective-prediction problem already solved upstream, and solving it again inside the model is neither necessary nor, on this evidence, effective.

For developers building assistive systems on neural decoders, decoder confidence should be carried into the action layer as an enforced admission decision rather than rendered into a prompt. Where the threshold belongs is harder, because a refusal is not free: in the repeated-attempt replay it costs the user another attempt, and enforcement improved tasks completed per attempt in two of five models while making it worse in one. The operating point therefore depends on the task’s severity and the retry burden the user will accept. For evaluators, unfaithful execution should be reported at matched coverage, since a bare rate is minimised by an agent that never acts. For regulators, the auditable object is the gate and its operating point, both specifiable and testable without reference to any model.

This study has several limitations. First, it was computational and enrolled no prospective participants, so we cannot know how a user would respond to an agent that declines to act, nor evaluate clarification behaviour. The repeated-attempt replay quantifies the mechanical cost of a refusal but not the human one, and its retries are population-level, which makes completion optimistic for a user whose decoding is persistently poor. Second, the confidence gated on was reconstructed from calibration recordings rather than read from an online decoder, because the corpus records no posterior; the calibration reported above bounds what any arm could have achieved. Third, ground truth was transmission fidelity rather than participant intent, since participants were copy-spelling prescribed strings, so an agent that correctly executes a faithfully transmitted but unintended request is scored as faithful. Fourth, *claude-sonnet-5* ran on a smaller, cost-driven 500-episode subset, so its intervals are wider. Fifth, all episodes came from one public corpus and one BCI paradigm, restricted to its four amyotrophic lateral sclerosis studies. Sixth, no provider seed was available, so generation is not reproducible run to run; a repeated-generation check is in the supplement. Seventh, model versions change, and these findings describe the identifiers queried between 28 and 31 August and on 9 and 10 September 2026.

Three studies follow. A prospective evaluation with people who use augmentative communication would test whether an enforced gate is acceptable to its users. A replication on a BCI exposing an online posterior would separate the ceiling imposed by reconstructed confidence from that imposed by the action layer. A replication in the able-bodied stratum of the same corpus, which holds 224 further participants, would show whether the pattern is specific to this population.

Placing an AI agent downstream of a neural decoder converts decoding errors into executed actions, and supplying that agent with calibrated decoder uncertainty as prompt text did not reduce unfaithful execution beyond what a threshold on the same number achieved at the same coverage. Semantic inference and uncertainty control are separable: the model can widen what an agent reaches, while the interface and its operating point keep the admission decision, specifiable and auditable while the model behind them changes.

## Methods

### Study Design

This was a computational benchmark study of an agentic large language model (LLM) acting on the output of a real P300 brain-computer interface (BCI) speller; no human participants were recruited and no system was deployed. The unit of analysis was the episode: three consecutive character selections emitted by the BCI decoder, together with the agent’s action in response.

The design was a 2 x 3 factorial (uncertainty source: none, self-confidence, or reconstructed decoder confidence; control mechanism: advisory, rendered into the prompt with the model’s own decision as the outcome, or enforced, withheld with admission decided by the harness) crossed with four reference arms: a deterministic non-LLM threshold gate, a random gate at matched coverage, an error-indicator arm bounding detection rather than correction, and a single-shot arm without a multi-turn loop, plus a 12-wording caution battery and 3 prompt renderings as an exposure and a nuisance factor. The full design comprised 34 cells, 32 involving an LLM (eMethods S1, eTable 1b); the fair-information comparison below is a separate, non-factorial set of five arms over the same 200 naturalistic episodes.

### When a Downstream Agent Can Improve on a Threshold

Let ***Γ*** ∈ {0, 1} denote whether a candidate action is faithful to the source command, and let

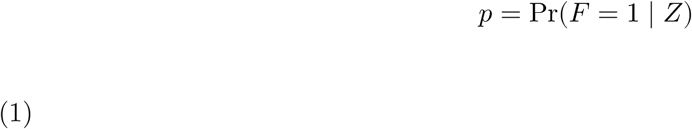

where *Z* is the reconstructed calibrated decoder confidence, estimating transmission correctness rather than action fidelity directly, since a mis-decoded string occasionally hashes to the same action as the correct one (88.5% of error-bearing episodes change the entailed action). Supplementary Note 1 shows the reconstructed confidence is a monotone transform of *p* under a fixed collision probability, an assumption no comparison here needs to hold exactly.

Executing an unfaithful action incurs cost *c* > 0; declining incurs retry cost *τ* ≥ 0, quantified below. The expected losses are

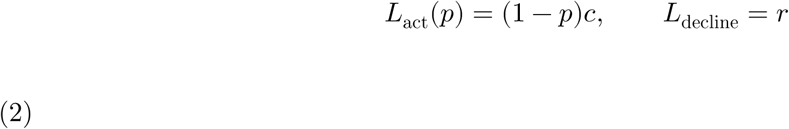

so the loss-minimising policy admits whenever (1 − *p*)*c* ≤ *τ*, equivalently

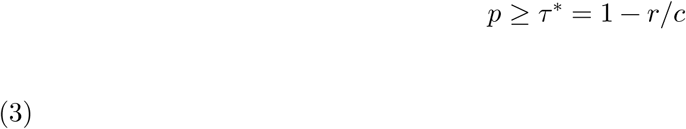

a threshold rule on *p* alone; the deterministic gate is the empirical analogue, thresholding the reconstructed decoder-confidence score.

A downstream agent can only improve on that policy using information the gate lacks: a better fidelity estimate than *p*, or a better cost estimate. If the agent observes additional information *H* beyond *Z*,

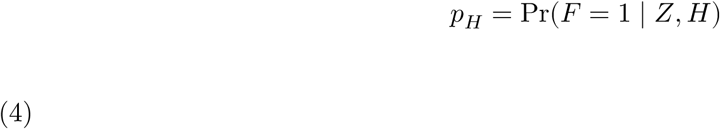

and, with *c* held fixed, the agent improves on the threshold policy only when *H* carries residual fidelity information *Z* does not:

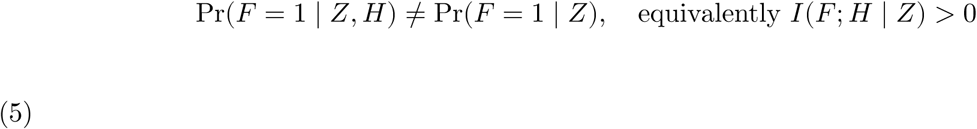

Holding *c* fixed is what the primary outcome does; where costs differ by action, an agent estimating context-specific cost can improve even when *I*(***F*** ; *H* ∣ *Z*) = 0, the setting the severity-aware endpoint addresses, while equation (5) is operative everywhere else.

The primary benchmark’s frozen codebook maps a decoded string to an action by a salted cryptographic hash, removing any semantic correspondence between string and action, so an agent tested there can only win by using *p* better than a threshold does. A semantically structured vocabulary reopens a legitimate second channel: a model that recognises a corrupted command from its meaning has fidelity information *p* does not encode and a threshold-only gate cannot use, exactly the manipulation that can make *I*(***F*** ; *H* ∣ *Z*) > 0, which the semantic benchmark below tests.

### Design Vocabulary

An **episode** is one fixed-length sequence of three consecutive character selections; a **cell** is one experimental condition (34 in the factorial design); a **scaffold** is one of three surface renderings of identical prompt content; an **arm** is the set of cells sharing an uncertainty source and control mechanism, pooled across scaffolds; an **episode run** is one execution of one episode in one cell by one model. Pooling across scaffolds therefore pools three runs of the same episode, and the participant-clustered bootstrap keeps every run from a sampled participant together. A count taken across the panel is reported as a number of model-by-arm or model-by-cell combinations; a result naming one model refers to that model’s runs of the arm or cell in question. Full cell and arm definitions, including how the caution and single-shot cells are kept from being pooled into the advisory baseline, are in eMethods S1.

### Data Source and Cohort

Episodes were constructed from bigP3BCI, a public P300 speller corpus, restricted to the four studies recorded in participants with amyotrophic lateral sclerosis (Studies B, F, L, and N);[13,14] the remaining studies, in able-bodied participants, were not analysed. The stratum contained 3,395 online selections from 47 participants across 9 sessions, accuracy 0.816 (per-participant range 0.219-1.000).

Episodes were formed by partitioning each test file into non-overlapping blocks of three selections, retained only with a non-null decoder score and correctness label on every selection; out-of-alphabet grid codes were dropped and counted. This produced 1,084 usable episodes, 364 (33.6%) with at least one decoding error.

The primary set is 1,065 primary-eligible episodes (usable pool minus 19 whose calibration fit came from an earlier session, a pre-specified exclusion), error prevalence 0.341. A second, exploratory set of 100 episodes was drawn earlier, balanced on consequence tier and enriched to 49 error-bearing episodes from 42 participants; it carries the 34-cell factorial and is labelled exploratory throughout. Full accounting is in eMethods S1 and eTable 1a.

### Reconstructed Decoder Confidence

bigP3BCI does not record an online posterior; the 36 grid-cell channels are binary stimulus-flash indicators rather than score accumulators. Confidence was therefore reconstructed from the calibration EEG of the same participant, session, and condition where available, referred to throughout as reconstructed calibrated decoder confidence, never as a posterior. Scores were mapped to probabilities with isotonic regression using out-of-fold predictions from grouped 5-fold cross-validation (participant as the grouping variable, so no participant contributed to both fitting and evaluation of any calibration statistic); episode-level confidence was the product of the three constituent selections’ calibrated confidences.

Calibration was assessed per study with expected calibration error and the Brier score (a pooled figure, reported in the Supplement, is not primary since opposite-signed bins cancel); transport was assessed by fitting on one study and evaluating on each other. Episodes whose calibration fit came from an earlier session were excluded from the primary analysis and retained for sensitivity analysis, an exclusion fixed before the LLM runs. Full calibration diagnostics are in eMethods S4.

### Agent, Tools, and Ground Truth

The action layer was a multi-turn tool-calling agent with four typed tools: read_buffer (returns the decoded string), lookup_action (resolves a string to its entailed action without terminating the episode), execute (takes one of nine permitted actions and terminates the episode), and abstain (terminates without acting). The environment held only the decoded string, so lookup_action returned the wrong action for a mis-decoded string rather than repairing it. The nine actions spanned three consequence tiers, presented in a frozen shuffled order carrying no tier grouping or tier-label text. On the analysed pool, 94 error-bearing episodes entailed an unrequested tier 3 action, and no error-free episode changed its entailed action. Schemas were identical across all arms and models; full schemas and the SHA-256-derived codebook are in eMethods S3.

Ground truth was transmission fidelity, not participant intent: an episode was scored faithful iff the executed action A equalled the action entailed by S, the string the channel carried (not S’, the string the decoder emitted). Participants were copy-spelling prescribed strings, so no claim was made about what any participant intended.

### Models and Inference Parameters

Five current-generation models were queried through OpenRouter between 28 and 31 August 2026, one per vendor: openai/gpt-5.6-luna, anthropic/claude-sonnet-5, google/gemini-3.7-flash, z-ai/glm-5.3-flash, and deepseek/deepseek-v4-flash. The fair-information comparison below widened this panel to ten under the same rule. anthropic/claude-sonnet-5 ran a frozen 500-episode subset of the primary pool rather than all 1,065, because its measured cost exceeded the prespecified projection; pooled cross-model estimates therefore use only the episode set common to every model, never raw pooling across differently-sized panels, with full subset provenance in eMethods S1.

Every request pinned a single provider endpoint by its routable tag with allow_fallbacks false, verified against the served provider on every response, since OpenRouter otherwise load-balances a model identifier across providers serving different quantizations. Temperature, top_p, and max_tokens were transmitted explicitly (0.7, 1.0, 512). Each episode was run once per cell per model, since the design is fully paired and repetition estimates stochasticity rather than adding neural observations; a repeated-generation check on two arms is in the Supplement. No random seed was set at the provider, and no claim of determinism is made. All 141,879 requests were served by the pinned provider; rate limiting, retry counts, and prompt digests are in eMethods S1, S5, and S2.

### Semantic Fair-Information Comparison

This post-hoc experiment, added after the primary results were obtained, answers the concern that the frozen hashed codebook structurally denies an agent any semantic information a threshold could not also use. It reused the 200 naturalistic episodes at their realized 0.34 error prevalence, producing 6,400 episode runs (US $19.46) across a panel widened from five models to ten. Full panel, endpoints, and run totals are in eMethods S11.

Three arms carried a language model: fair:llm_vocab:advisory (unchanged agent loop, all nine commands disclosed, confidence rendered into the prompt: the information-symmetric counterpart of the primary benchmark’s advisory cell); fair:llm_vocab:enforced (same loop and vocabulary, confidence withheld, proposals swept across a threshold grid); and fair:hybrid_semantic_gate (a single-call harness with no tools or history, in which the model proposes a semantic correction as text and a deterministic threshold outside the model decides admission, confidence withheld so the architecture never reintroduces it through the prompt).

Writing ℎ*_θ_*(*S*^′^, *H*) for the model’s proposed correction given the decoded string and disclosed vocabulary, *s* for the recalibrated out-of-fold score an arm thresholds (ordering episodes approximately as *p* does, per Supplementary Note 1), and *τ* for that threshold, the hybrid architecture is

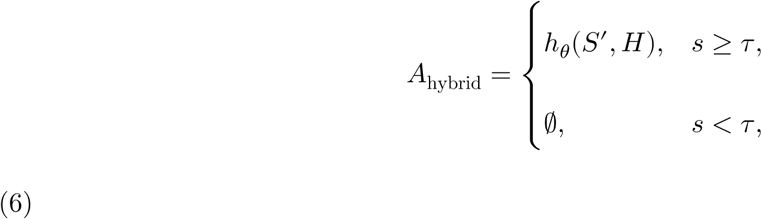

the same threshold rule as equation (3), now admitting a semantic correction: ℎ*_θ_* supplies the information *H* equation (5) shows an agent can legitimately contribute, while *s* ≥ *τ* retains admission outside the model. This reuses equation (3)’s threshold FORM, not its optimality proof, which holds only for the fidelity of the action the gate is asked to admit, not the model’s proposed correction, whose fidelity *s* was never calibrated against; whether this score is nonetheless workable for a semantically corrected action is what the Results test.

Two comparators carried no model: fair:exact_resolver_gate (exact-match codebook lookup) and fair:lexical_resolver_gate (edit-distance resolution, max distance 2), paired with the same gate. The lexical resolver is the primary comparator throughout this paper and the one that motivated this experiment, handed the nine command strings literally, so here the models are too. Both the advisory arm’s confidence and every gate here use the out-of-fold isotonic score rather than the primary benchmark’s per-selection product score, so the comparison stays information-symmetric; full detail is in eMethods S11.

### Agent Harness

The loop ran for at most 6 turns, terminating when the model called execute or abstain, when a response contained no parseable tool call, or when the turn limit was reached. read_buffer was not mandatory; lookup_action was non-terminal and could be called repeatedly. Every tool call in a response was executed in the order returned. A response with no valid tool call, malformed structure, or invalid JSON arguments was recorded as a parse failure and ended the episode; an episode reaching the turn limit without a terminal call was scored as uncovered rather than dropped. Invalid enumeration values were never repaired or retried. Full termination, parsing, and retry rules are in eMethods S3.

### Self-Confidence Elicitation

Under advisory control the self-confidence arm added one sentence to the system prompt asking the model to state its confidence that it understood the message, 0 to 1; the value was never read back, calibrated, or used to gate anything, so this arm tests what asking a model to self-assess buys, not what a well-calibrated self-assessment would buy. Under enforced control the prompt carried no uncertainty text, so the self-confidence and decoder-confidence enforced cells were identical.

### Model Panel Selection

The panel was fixed before any experimental run, from the live OpenRouter model list as it stood on 28 August 2026: one current-generation model per vendor, chosen to span the list’s price range (cheapest capable, most expensive as a ceiling arm, one mid-priced, two open-weight), with prices read from the live list rather than recalled. The panel is a prespecified selection, not a random sample, reflecting the cost constraint an assistive deployment would face rather than a claim to cover frontier reasoning capability.

### Outcomes

The primary outcome was **unfaithful execution** at matched action coverage: the proportion of executed actions differing from the action the source string entails. The term is deliberately not “unsafe execution”: the primary outcome weights every error equally, while **unsafe execution**, reported alongside, is the severity-aware endpoint, execution of an unrequested tier 3 action. Coverage was the proportion of episodes on which the agent executed any action; a bare unfaithful-execution rate was not used as primary, since an agent that never acts minimises it.

For a policy admitting whenever *s* ≥ *τ* (*s* need not be a calibrated probability for arms that threshold something other than the decoder confidence), coverage and selective risk are

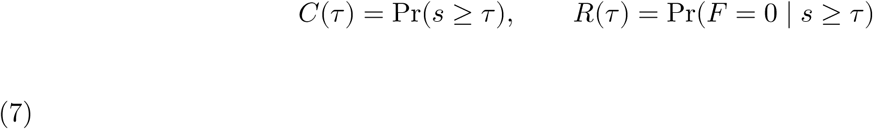

and two policies are compared at matched coverage *C* by

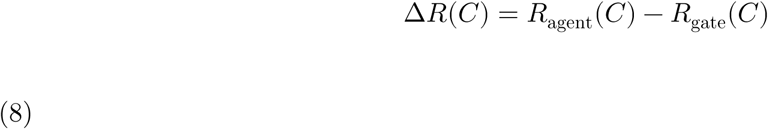

where Δ*R*(*C*) < 0 indicates the agent improved on deterministic admission. Advisory arms, having no continuous knob, enter as a fixed point tested against the gate’s swept curve at that coverage; continuous-knob arms were swept to trace a full risk-coverage curve, summarised by the area above a coverage floor of 0.10 over the shared support. Secondary outcomes were parse-failure rate, abstention mechanism, consequence-tier stratification, the caution battery, and calibration. Full procedures are in eMethods S6.

### Statistical Analysis

Primary analyses included all primary-eligible episodes per model; analyses restricted to error-bearing episodes were secondary. Risk differences were estimated as differences in unfaithful- execution proportions, with 95% confidence intervals from a participant-clustered bootstrap (2,000 resamples, seed 20260828), each replicate drawing one joint participant sample applied to both arms, since independent resampling would discard the paired design. Clustering on participant applied throughout (47 primary, 42 exploratory participants). Risk ratios use the Haldane-Anscombe correction. P values came from the bootstrap intervals, adjusted by Benjamini-Hochberg within two pre-specified families: per-model uncertainty-source contrasts and the 12-wording caution battery. Significance threshold was *P* < .05.

A cell whose parse-failure rate exceeded 15%, fixed before the runs, is labelled rather than removed, since a cell that fails to emit a valid tool call is showing a behaviour the design should retain; labelled cells are in Results and eTable 4, with a sensitivity sweep removing them progressively. Software versions and a cross-interpreter reproducibility check are in eMethods S6.

### Repeated-Attempt Empirical Replay

The primary analyses score a single decision per episode, so an abstention is charged as coverage lost; a person whose speller declines has not finished the task and must attempt again. To quantify that cost, the 200 naturalistic episodes were replayed as repeated-attempt tasks in a persistent assistive-action sandbox where each action produces an inspectable state change, under a protocol fixed before any replay was run.

A task is one intended command. On each attempt a donor episode for that command is drawn uniformly without replacement, and the policy’s recorded decision determines the outcome: faithful execution completes the task, unfaithful execution ends it wrongly, a decline consumes an attempt, up to three. Wrong execution is terminal, since no human correction is simulated. Every policy was evaluated over the identical donor-draw distribution, so contrasts are paired; because every decision was already recorded, the outcome distribution was computed exactly by enumerating the draw tree. All seventeen policies are in eTable 10.

This is a system-level replay, not validation of an interactive assistive system: the agent is memory- less across attempts and the replay is population- rather than participant-specific. Full methodology is in eTable 10’s methods note.

### Standard Protocol Approvals, Registrations, and Patient Consents

This study analysed a fully de-identified, publicly available BCI corpus and enrolled no participants, so institutional review board approval and informed consent were not required. No clinical intervention was performed and no trial registration applies. Reporting follows MI-CLAIM for clinical artificial intelligence studies. Analysis code and the complete run manifest, including the frozen episode identifiers, prompt set, tool schemas, and action codebook with their SHA-256 digests, are available at https://github.com/BRIDGE-GenAI-Lab/AI-Agents-in-BCIs.

## Supporting information

full appednix

## Data Availability

All data produced are available online at https://github.com/BRIDGE-GenAI-Lab/AI-Agents-in-BCIs

## References

1. Farwell LA, Donchin E. Talking off the top of your head: toward a mental prosthesis utilizing event-related brain potentials. Electroencephalogr Clin Neurophysiol. 1988;70(6):510–523. doi:10.1016/0013-4694(88)90149-6

2. Wolpaw JR, Birbaumer N, McFarland DJ, Pfurtscheller G, Vaughan TM. Brain-computer interfaces for communication and control. Clin Neurophysiol. 2002;113(6):767–791. doi:10.1016/S1388-2457(02)00057-3

3. Mainsah B, Fleeting C, Balmat T, Sellers E, Collins L. bigP3BCI: an open, diverse and machine learning ready P300-based brain-computer interface dataset. Version 1.0.0. PhysioNet; 2025. doi:10.13026/0byy-ry86

4. Willett FR, Kunz EM, Fan C, et al. A high-performance speech neuroprosthesis. Nature. 2023;620(7976):1031–1036. doi:10.1038/s41586-023-06377-x

5. Metzger SL, Littlejohn KT, Silva AB, et al. A high-performance neuroprosthesis for speech decoding and avatar control. Nature. 2023;620(7976):1037–1046. doi:10.1038/s41586-023-06443-4

6. Javdani S, Admoni H, Pellegrinelli S, Srinivasa SS, Bagnell JA. Shared autonomy via hindsight optimization for teleoperation and teaming. Int J Rob Res. 2018;37(7):717–742. doi:10.1177/0278364918776060

7. Pinegger A, Faller J, Halder S, Wriessnegger SC, Muller-Putz GR. Control or non-control state: that is the question! An asynchronous visual P300-based BCI approach. J Neural Eng. 2015;12(1):014001. doi:10.1088/1741-2560/12/1/014001

8. Chow CK. On optimum recognition error and reject tradeoff. IEEE Trans Inf Theory. 1970;16(1):41–46. doi:10.1109/TIT.1970.1054406

9. Geifman Y, El-Yaniv R. Selective classification for deep neural networks. In: Advances in Neural Information Processing Systems 30 (NIPS 2017). Curran Associates; 2017:4878–4887.

10. Andriushchenko M, Souly A, Dziemian M, et al. AgentHarm: a benchmark for measuring harmfulness of LLM agents. In: The Thirteenth International Conference on Learning Representations (ICLR 2025); 2025.

11. Ruan Y, Dong H, Wang A, et al. Identifying the risks of LM agents with an LM-emulated sandbox. In: The Twelfth International Conference on Learning Representations (ICLR 2024); 2024.

12. Vansteensel MJ, Pels EGM, Bleichner MG, et al. Fully implanted brain-computer interface in a locked-in patient with ALS. N Engl J Med. 2016;375(21):2060–2066. doi:10.1056/NEJMoa1608085

13. Sellers EW, Donchin E. A P300-based brain-computer interface: initial tests by ALS patients. Clin Neurophysiol. 2006;117(3):538–548. doi:10.1016/j.clinph.2005.06.027

14. Nijboer F, Sellers EW, Mellinger J, et al. A P300-based brain-computer interface for people with amyotrophic lateral sclerosis. Clin Neurophysiol. 2008;119(8):1909–1916. doi:10.1016/j.clinph.2008.03.034

15. Gorenshtein A, Omar M, Barash Y, Nadkarni GN, Klang E. Large language models integrated into brain-computer interfaces for communication and control: a systematic review. Biomed Phys Eng Express. 2026;12(3):035077. doi:10.1088/2057-1976/ae737b

16. Lee JY, Lee S, Mishra A, et al. Brain-computer interface control with artificial intelligence copilots. Nat Mach Intell. 2025;7(9):1510–1523. doi:10.1038/s42256-025-01090-y

17. Gorenshtein A, Omar M, Glicksberg BS, Nadkarni GN, Klang E. AI Agents in Clinical Medicine: A Systematic Review. medRxiv. 2025. doi:10.1101/2025.08.22.25334232

18. Qiu J, Lam K, Li G, Acharya A, Wong TY, Darzi A, Yuan W, Topol EJ. LLM-based agentic systems in medicine and healthcare. Nat Mach Intell. 2024;6(12):1418–1420. doi:10.1038/s42256-024-00944-1

19. Kumaran D, Daw N, Osindero S, Velickovic P, Patraucean V. Causal evidence that language models use confidence to drive behaviour. Nat Mach Intell. Published online September 7, 2026. doi:10.1038/s42256-026-01293-x

